# Cross-sectional and longitudinal associations of smoking behavior with central arterial hemodynamic measures: The Framingham Heart Study

**DOI:** 10.1101/2024.06.30.24309741

**Authors:** Leroy L. Cooper, Sana Majid, Na Wang, Jessica L. Fetterman, Joseph N. Palmisano, Emelia J. Benjamin, Ramachandran S. Vasan, Gary F. Mitchell, Naomi M. Hamburg

**Author notes:** Corresponding author: (LLC).

## Abstract

**Background:** Cigarette smoking is a leading modifiable cardiovascular disease risk factor. Cross-sectional studies demonstrated variable relations of smoking behavior with measures of vascular function. Additionally, the impact of persistent versus changes in smoking behaviors on alterations in central arterial function is unclear.

**Methods and Results:** We assessed associations of smoking behavior and intensity with measures of central arterial hemodynamics in 6597 participants (51.5% never smoked, 34.8% formerly quit, 4.3% recent quit, and 9.3% currently smoking) of the Framingham Heart Study (N=3606 [55%] women). In cross-sectional models, central vascular measures were different across categories of smoking behavior. For example, augmentation index was higher among participants who formerly quit smoking (least squares mean±standard error=14.1±0.4%; *P*<0.001) and were currently smoking (18.1±0.5%; *P*<0.001) compared to participants who never smoked (12.6±0.3%). Among participants who were currently smoking, higher cigarettes per day (estimated *B*=1.41; 95% CI, 0.47—2.34; *P*=0.003) was associated with higher augmentation index. Among participants who had quit smoking, higher pack-years was associated with higher augmentation index (est. *B*=0.85; 95% CI, 0.60—1.14) and central pulse pressure (est. *B*=0.84; 95% CI, 0.46—1.21). Using restricted cubic splines, we observed a negative linear association for augmentation index, but a distinct nonlinear association for characteristic impedance and central pulse pressure, with higher time since quit (all *P*<0.001). In longitudinal models, we observed higher increases in augmentation index among participants who persistently quit (4.62±0.41%; *P*<0.001) and persistently smoked (5.48±0.70%; *P*=0.002) compared to participants who persistently never smoked (3.45±0.37%).

**Conclusion:** Central arterial measures are sensitive to smoking status and intensity and changes in smoking behavior. Longer smoking cessation may partially revert central arterial measures to levels comparable to those with lower smoking exposure.

## Introduction

Cigarette smoking is a leading modifiable risk factor for cardiovascular disease (CVD) and contributes to nearly half a million deaths annually in the United States [1, 2]. Smoking duration and intensity are known to contribute to the toxicologic burden of CVD and increase disease risk [3–5]. For example, smoking contributes to a range of cardiovascular consequences, including endothelial dysfunction and inflammation, as well as conditions such as coronary artery disease, myocardial infarction, heart failure, stroke, aneurysms, and peripheral vascular disease [6]. Conversely, smoking cessation is associated with reduction of morbidity and mortality and improvement to cardiovascular health [7–9]. Measures of vascular central hemodynamics and aortic stiffness are preclinical predictors of CVD events [10–14]. Several cross-sectional studies have shown inconsistent associations of smoking status and intensity with vascular hemodynamic measures that contribute to CVD that were assessed in various vascular beds [15–22]. Additionally, the impact of changes in smoking behaviors on central arterial hemodynamic measures have yet to be elucidated in well-characterized cohorts with longitudinal tracking of smoking status and comprehensive vascular hemodynamics. Thus, the putative effects of continuous and changes in smoking behavior (intensity, cessation, and relapse) on central arterial hemodynamics are not defined. We aimed to assess the cross-sectional and longitudinal associations of smoking behavior with measures of central arterial hemodynamics in samples of the Framingham Heart Study (FHS).

## Materials and Methods

Our study followed the Strengthening the Reporting of Observational Studies in Epidemiology (STROBE) reporting guidelines [23]. The procedures for requesting data from the Framingham Heart Study can be found at https://www.framinghamheartstudy.org/.

### Study Sample

The study samples were drawn from the Framingham FHS Offspring, New Offspring Spouses, Generation 3, Omni 1, and Omni 2 cohorts, which have been described in detail previously [24–26]. Participants who attended examinations when both smoking data and measures of vascular function were reliably collected for the index examination visit and a follow-up visit were eligible for this investigation (examinations 8 [08-03-2005 to 25-01-2008] and 9 [11-04-2011 to 24-04-2014] for Offspring; examinations 2 [27-05-2008 to 14-03-2011] and 3 [11-04-2016 to 30-04-2019] for New Offspring Spouses, Generation 3, and Omni 2; and examinations 3 [02-02-2007 to 10-01-2008] and 4 [24-08-2011 to 24-03-2014] for Omni 1). 7119 participants were eligible for this investigation at the index examination visit. We excluded 522 due to missing data or a quit history of less than 1 year (among participants who reported quitting) to censor for unsuccessful quit attempts and relapses. Three participants with inconsistent self-reported smoking behavior were classified among those with missing smoking data. At follow-up, 1243 participants were further excluded due to loss to follow-up, death, or missing smoking data. All protocols were approved by the Boston University Medical Center’s Institutional Review Board, and all participants provided written informed consent.

### Smoking Status and Intensity

At each examination, participants were categorized by smoking status based on their responses to a self-report questionnaire. We defined currently smoking as smoking regularly within 12 months of each respective examination. We defined smoking cessation (or quitting) behavior for participants who 1) responded “no” when asked if they smoke cigarettes regularly in the last year; 2) responded “no” when asked if they were currently smoking (as of 1 month ago); 3) indicated current or regular smoking at a prior visit; and 4) provided an age if they have stopped smoking completely. To better capture the effects of smoking cessation, we further categorized participants by their quit history as recently quit (participants who reported regular cigarette use at prior visits but had quit by the index examination visit) and formerly quit (participants who reported no regular cigarette use at or before the visit prior to the index examination visit) based on responses given for the questionnaire. We defined “never smoked” when participants reported never having smoked. Participants who were currently smoking and those who quit were asked their age when they started smoking, cigarettes smoked per day when smoking or when they smoked, and age at quitting (if smoking ceased). For these questions, we assessed smoking duration, cumulative pack years, and years since quitting. Using the same questionnaire, we assessed change in smoking status between the two exam cycles for longitudinal analyses.

### Hemodynamic assessment with arterial tonometry

Hemodynamic assessment with arterial tonometry and pulsed doppler electrocardiography was performed as previously described [27, 28]. We obtained non-invasive arterial tonometry with simultaneous electrocardiography from supine participants for the brachial, radial, femoral, and carotid arteries using a custom tonometer. During the tonometric assessment of the carotid artery, we performed pulsed Doppler of the left ventricular outflow tract to assess aortic flow. We digitized and transferred tonometric data and Doppler electrocardiography data to a core laboratory (Cardiovascular Engineering, Inc., Norwood, MA) for blinded analyses. We signal-averaged and synchronized tonometry waveforms using the electrocardiographic R-wave and then calculated mean arterial pressure (MAP) as the integral of the signal-averaged brachial pressure tonometry waveform [28]. We estimated pulse wave velocities from tonometry waveforms and body surface measurements that adjusted for parallel transmission in the aortic arch and brachiocephalic artery as previously described [29]. We calculated the carotid-femoral pulse wave velocity (CFPWV), carotid-brachial pulse wave velocity, and carotid-radial pulse wave velocity as the ratios of the adjusted transit distance and the pulse transit time difference between carotid and the femoral, brachial, and radial sites, respectively. We calculated the pulse wave velocity ratio as the CFPWV divided by the carotid-brachial pulse wave velocity. We calculated central pulse pressure as the difference between carotid systolic and diastolic blood pressures. We defined forward pressure wave amplitude (FWA) as the difference between pressure at the foot and at the peak of the forward pressure waveform by performing time domain wave separation analysis using central pressure and flow [30]. The global reflection coefficient was defined as backward wave amplitude divided by FWA. We calculated augmentation index (AI) as the fraction of central pulse pressure attributable to late systolic pressure. Characteristic impedance was calculated in the time domain as the ratio of the pressure increase and the flow increase during the time interval between flow onset and 95% of peak flow [30].

### Clinical evaluation and covariates

Medical history was acquired, and a physical examination was performed routinely at each examination visit. Age, sex, use of antihypertensive and hyperlipidemia medications, and CVD history were assessed through questionnaires. Height (meters) and weight (kilograms) were assessed during the examination. Body mass index was calculated as weight in kilograms divided by height in meters squared. Heart rate and blood pressures were assessed during tonometry. Serum cholesterol levels were measured from fasting blood test. Criteria for diabetes were a fasting glucose level of ≥126 mg/dL (7.0 mmol/L) or treatment with insulin or an oral hypoglycemic agent.

### Statistical analysis

We tabulated characteristics for the study sample. CFPWV was inverted to limit heteroscedasticity, then multiplied by −1000 to convert units to ms/m and rectify the directionality of associations with aortic stiffness. We selected covariates *a priori* as follows: age, age^2^, sex, cohort, body mass index, heart rate, mean arterial pressure, total to high-density lipoprotein cholesterol ratio, triglycerides, prevalent CVD, lipid-lowering medication use, hypertension treatment, and prevalent diabetes. The triglycerides variable was natural logarithmically transformed to normalize its skewed distribution. To maximize sample sizes, we excluded participants missing individual tonometry measures on an analysis-by-analysis basis.

In cross-sectional analyses, we used multivariable linear regression models to relate smoking status (currently smoking, formerly quit, recently quit, and never smoked) with central arterial hemodynamic measures. The never smoked group was the reference group. We estimated least squares means based on regression models with central arterial hemodynamic measures as dependent variables and smoking status groups as independent variables, adjusting for the above-mentioned covariates. In secondary analyses, we used multivariable linear regression models to relate smoking status with peripheral arterial hemodynamic measures. To assess the association of smoking intensity among participants who ever smoked, we used multivariable linear regression models to relate cigarettes per day (among participants who were currently smoking) and pack-years (among participants who had quit smoking) with central arterial measures. We estimated multivariable-adjusted nonlinear relations of time since quit with central arterial measures using restricted cubic splines.

For longitudinal analyses, we calculated change from the index examination visit to the follow-up exam in central arterial measures for each of the groups as follows: persistently never smoked, persistently quit, recently quit, and persistently smoking. Participants in a change in smoking status group with a low sample size were excluded from the analysis. We assessed the association of changes of central arterial measures with smoking status in the longitudinal sample using multivariable linear regression adjusted for the aforementioned covariates and central arterial hemodynamic values at the index examination visit. We estimated least squares means based on regression models with change in vascular measures as dependent variables and smoking status groups as independent variables, adjusting for the above-mentioned covariates. All analyses were performed with SAS version 9.4 for Windows (SAS Institute, Cary, NC). Bonferroni-adjusted, 2-sided *P* values were used to assess statistical significance.

## Results

We included 6597 participants (3606 [55%] women) in the analyses. A flow diagram for the analysis samples is presented in Fig 1, and clinical characteristics and arterial hemodynamic measures of the study participants stratified by smoking category at the index examination visit are presented in Table 1 and S1 Table, respectively. Participants with a shorter quit history (recently quit) had a median time since quit of 3.2 years, and participants with a longer quit history (formerly quit) had a median time since quit of 23.1 years.

**Fig 1.**
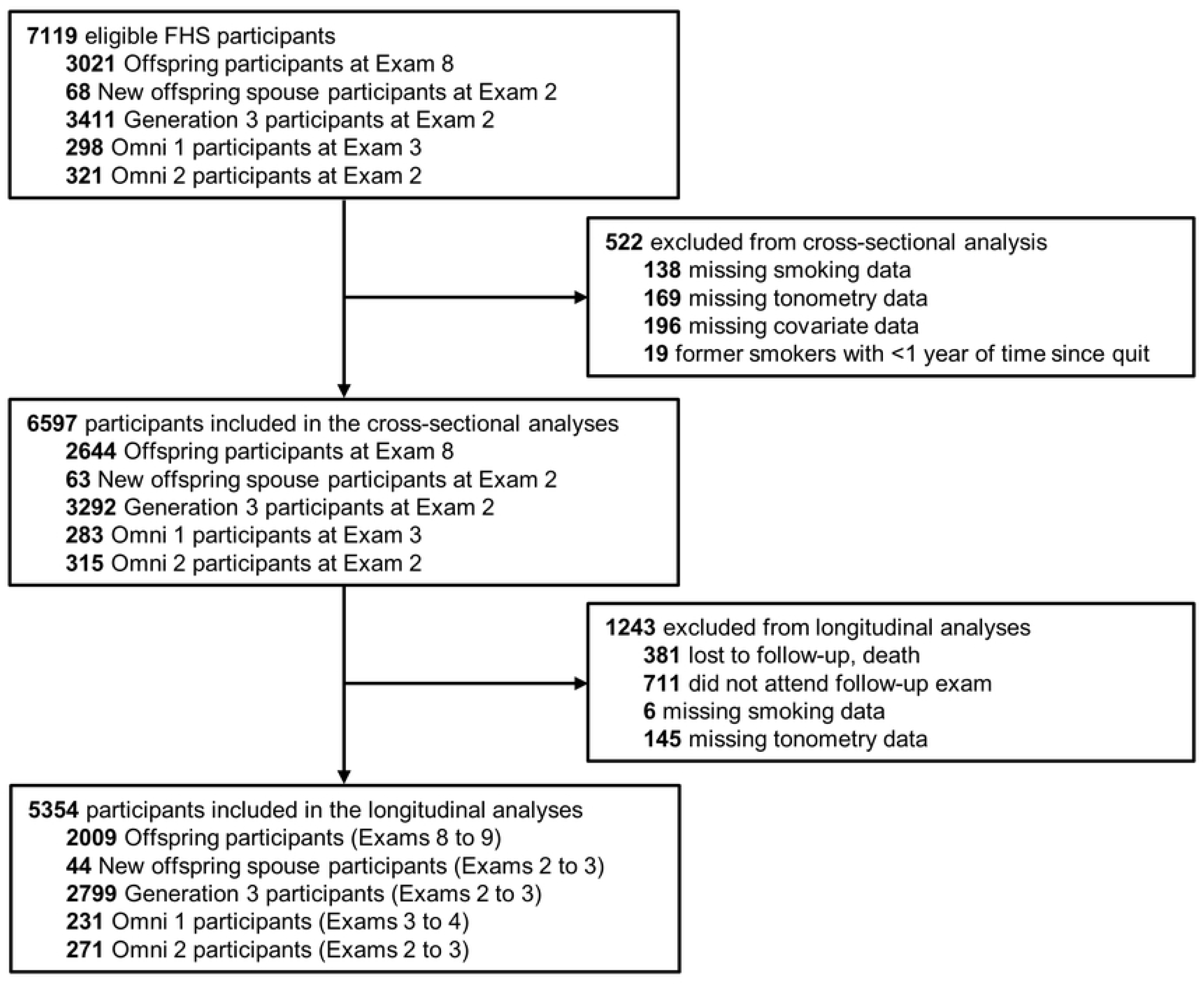
Flow diagram of the Framingham Heart Study cohorts comprising the analytic samples.

**Table 1.** Clinical characteristics at the index examination visit.

| **Variable** | **Overall**<br>**N=6597** | **Never**<br>**Smoked**<br>**N=3399** | **Formerly**<br>**Quit**<br>**N=2299** | **Recently**<br>**Quit**<br>**N=284** | **Currently**<br>**Smoking**<br>**N=615** |
| --- | --- | --- | --- | --- | --- |
| Age, y | 55.3±13.1 | 52.7±13.2 | 60.4±12.0 | 52.2±12.3 | 52.2±11.6 |
| Women, n (%) | 3606 (55) | 1888 (56) | 1246 (54) | 153 (54) | 319 (52) |
| Body mass index, kg/m<sup>2</sup> | 28.0±5.6 | 27.8±5.6 | 28.4±5.4 | 29.4± 5.7 | 27.6±6.0 |
| Heart rate, beats per minute | 63±10 | 62±10 | 62±10 | 64±10 | 66±10 |
| Mean arterial pressure, mm Hg | 92±13 | 91±13 | 94±12 | 92±13 | 91±12 |
| Total-to-HDL cholesterol ratio | 3.4±1.1 | 3.4±1.1 | 3.4±1.0 | 3.6±1.2 | 3.7±1.3 |
| Triglycerides, mg/dL | 97 [70, 136] | 94 [68, 131] | 98 [72, 137] | 106 [74, 159] | 104 [76, 147] |
| Prevalent cardiovascular disease, n (%) | 557 (8) | 174 (5) | 288 (13) | 36 (13) | 59 (10) |
| Lipid-lowering medications, n (%) | 1825 (28) | 765 (23) | 833 (36) | 77 (27) | 150 (24) |
| Prevalent hypertension, n (%) | 2502 (38)<sup>a</sup> | 1106 (33) | 1096 (48) | 102 (36) | 198 (32) |
| Hypertension treatment, n (%) | 2046 (31) | 899 (26) | 918 (40) | 78 (28) | 151 (25) |
| Prevalent diabetes, n (%) | 600 (9) | 250 (7) | 276 (12) | 29 (10) | 45 (7) |
| Age of smoking initiation, y | 17.7±3.8<sup>b</sup> | --- | 17.7±3.7 | 18.0±4.2 | 17.3±4.0 |
| Smoking duration, y | 21.0±13.5<sup>b</sup> | --- | 17.1±11.5 | 26.9±13.5 | 32.9±12.7 |
| Cumulative pack-years, y | 20.7±20.3<sup>b</sup> | --- | 17.6±18.3 | 22.0±22.7 | 31.7±22.4 |
| Years since quitting | 22.5±12.3<sup>c</sup> | --- | 24.8±11.1 | 4.2±3.3 | --- |
| Cigarettes per day, n | 13.5±9.4<sup>d</sup> | --- | --- | --- | 13.5±9.4 |

HDL, high-density lipoprotein. Values are mean±standard deviation, number (%), or median [25th, 75th percentile].
<sup>a</sup>N=6595.
<sup>b</sup>N ranges from 3058 to 3195.
<sup>c</sup>N=2582.
<sup>d</sup>N=612.

We present comparisons of least squares means estimates of central arterial hemodynamic measures according to smoking status at Visit 1 in Table 2. Compared to participants who never smoked, participants who formerly quit and were currently smoking had higher augmentation index. Participants who formerly quit had higher mean global reflection coefficient, and participants who were currently smoking had higher mean central pulse pressure, global reflection coefficient, and peak aortic flow rate compared to those who never smoked. We observed no significant cross-sectional differences for other central vascular measures by smoking status. We present comparisons of least squares means estimates of peripheral arterial hemodynamic measures according to smoking status at Visit 1 in S2 Table. Compared to participants who never smoked, participants who were currently smoking had lower carotid-brachial pulse wave velocity. We observed no significant cross-sectional differences for other peripheral vascular measures by smoking status.

**Table 2.** Estimated least squares means of central arterial hemodynamic measures according to smoking status at Visit 1.

| Outcome Variable | Never<br>Smoked | Formerly<br>Quit | Recently<br>Quit | Currently<br>Smoking |
| --- | --- | --- | --- | --- |
| Augmentation index, % | 12.6±0.3<br>(reference) | 14.1±0.4<br>($<0.001$) | 14.5±0.7<br>(0.003) | 18.1±0.5<br>($<0.001$) |
| niCFPWV, ms/m | -128±1<br>(reference) | -128±1<br>(0.58) | -127±1<br>(0.49) | -130±1 (0.05) |
| Central pulse pressure, mm Hg | 61.5±0.4<br>(reference) | 62.3±0.5<br>(0.02) | 63.4±0.9<br>(0.01) | 64.8±0.7<br>($<0.001$) |
| Forward pressure wave, mmHg | 51.8±0.4<br>(reference) | 51.9±0.4<br>(0.95) | 52.8±0.8<br>(0.17) | 52.3±0.6<br>(0.33) |
| Characteristic impedance, dyne•sec/cm<sup>5</sup> | 223±2<br>(reference) | 219±2<br>(0.04) | 216±4<br>(0.10) | 215±3 (0.01) |
| Global reflection coefficient, % | 36.1±0.2<br>(reference) | 36.7±0.2<br>($<0.001$) | 36.4±0.4<br>(0.49) | 38.1±0.3<br>($<0.001$) |
| Peak aortic flow rate, mL/sec | 307±2<br>(reference) | 308±2<br>(0.38) | 317±4<br>(0.004) | 316±3<br>($<0.001$) |

We present multivariable cross-sectional associations of central arterial hemodynamic measures with surrogates of smoking intensity in Table 3. Among participants who were currently smoking, higher cigarettes per day (estimated *B*=1.41; 95% CI, 0.47—2.34; *P*=0.003) was associated with higher augmentation index but was not associated with other central arterial measures. Among participants who had quit smoking, higher pack-years was associated with higher augmentation index (est. *B*=0.85; 95% CI, 0.60—1.14) and central pulse pressure (est. *B*=0.84; 95% CI, 0.46—1.21) but was not associated with other central arterial measures. Fig 2 depicts multivariable-adjusted restricted cubic splines plots of the relations of time since quit with select central arterial hemodynamic measures. We observed a steep, linear negative association for augmentation index (*P*<0.001) with higher time since quit. However, we observed distinct nonlinear associations of time since quit with characteristic impedance (*P*<0.001) and central pulse pressure (*P*<0.001). Among participants with shorter quit history, characteristic impedance was similar, but we observed progressively higher characteristic impedance after 25 years of smoking cessation. Among participants with shorter quit history, central pulse pressure was progressively lower until after 25 years of smoking cessation when central pulse pressure was progressively higher among those with longer quit history.

**Fig 2.**
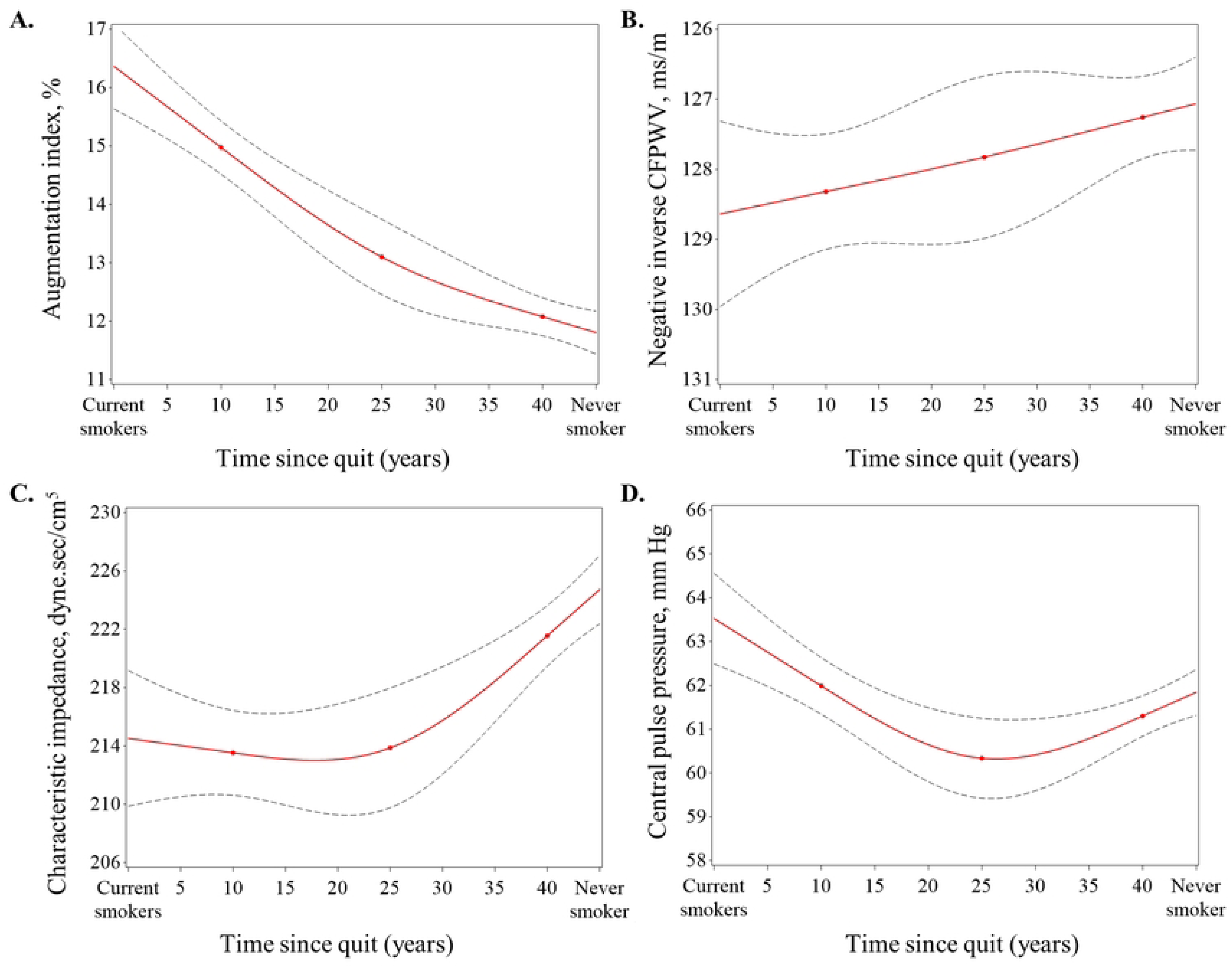
Multivariable-adjusted spline illustrates the associations of time since quit with central arterial hemodynamic measures. Restricted cubic splines (red solid lines) with 95% confidence intervals (dotted lines) derived from associating time since quit with (A) augmentation index, (B) negative inverse carotid-femoral pulse wave velocity (niCFPWV), (C) characteristic impedance, and (D) central pulse pressure. The *x*-axis ranges from currently smoking (minimum) to never smoked (maximum), and the values captured among participants who have quit based on time since smoking cessation. We placed knots at the 25^th^, 50^th^, and 100^th^ percentiles of the distribution of time since quit. All models were adjusted for age, age^2^, sex, cohort, body mass index, heart rate, mean arterial pressure, total to high-density lipoprotein cholesterol ratio, triglycerides, prevalent cardiovascular disease, lipid-lowering medication use, hypertension treatment, and prevalent diabetes. *P* for overall association is <0.001 for augmentation index; 0.07 for niCFPWV; <0.001 for characteristic impedance; and <0.001 for central pulse pressure. *P* for non-linearity is 0.08 for augmentation index; 0.93 for niCFPWV; and 0.02 for characteristic impedance; and <0.001 for central pulse pressure.

**Table 3.**
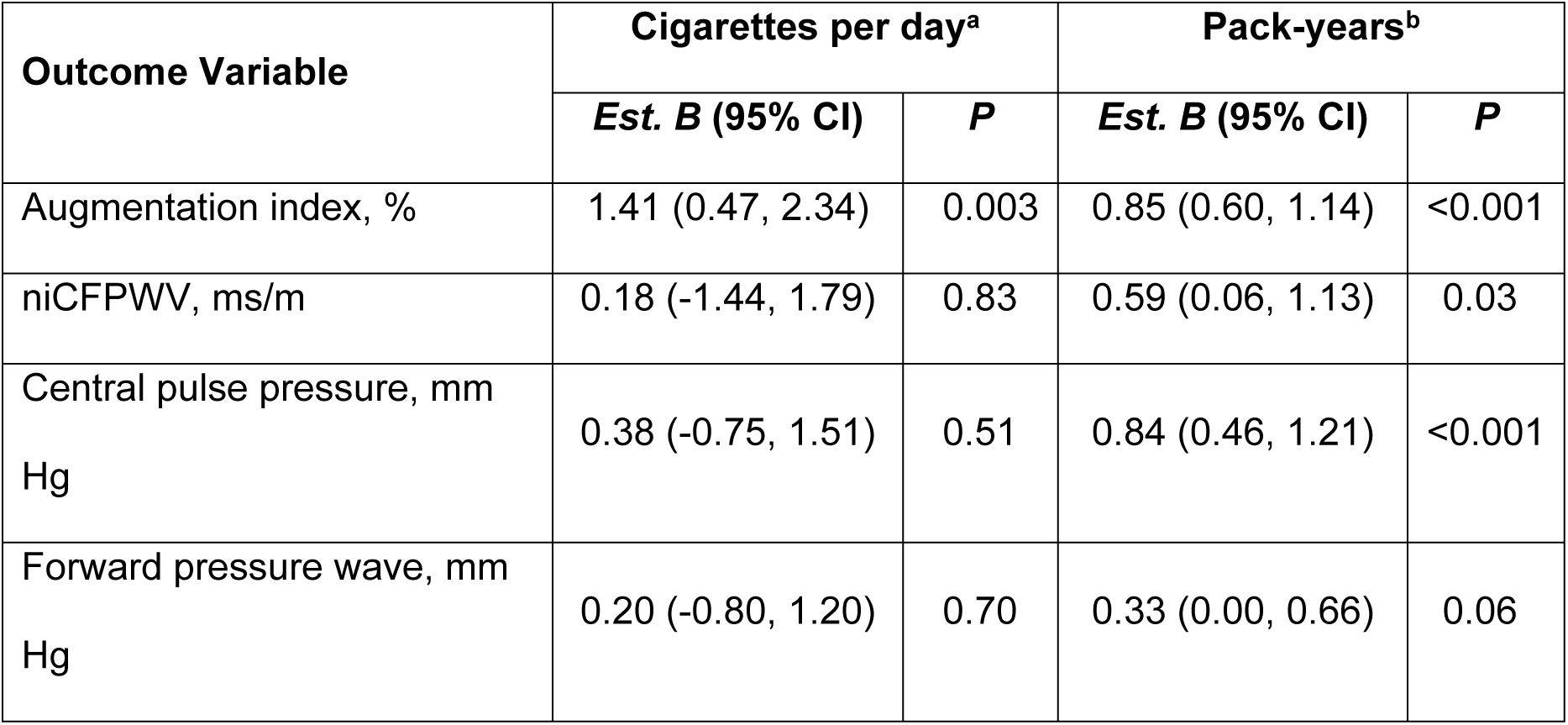
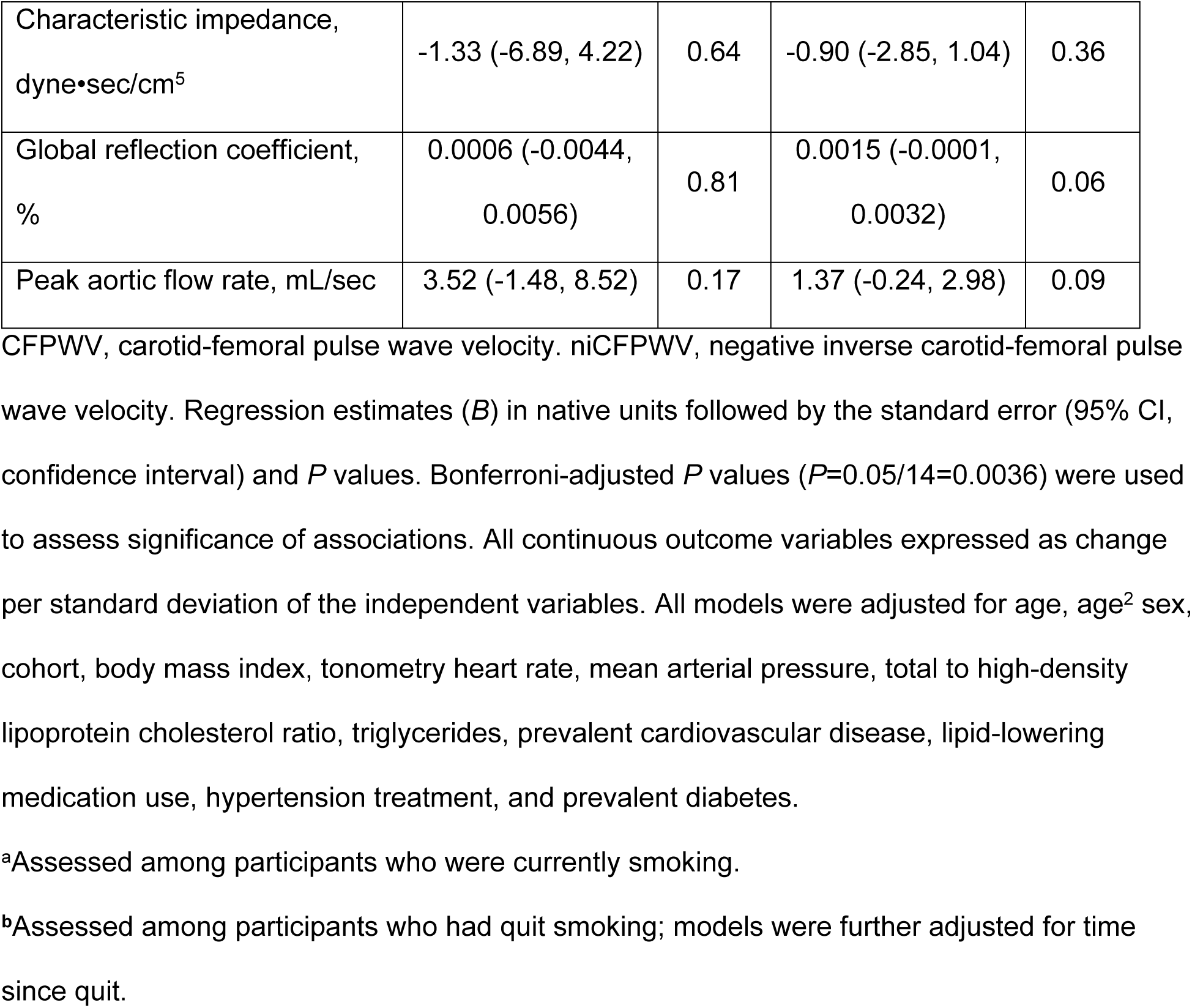
Multivariable cross-sectional associations of central arterial hemodynamic measures with smoking intensity at Visit 1.

We present a comparison of least squares means estimates of the longitudinal change in central arterial hemodynamic measures between two examination visits by smoking behavior in Table 4. We excluded participants who exhibited a change in smoking status with low sample sizes (never smoked to quit, never smoked to currently smoking, and quit to currently smoking). Compared to participants who persistently never smoked, participants who persistently quit and persistently smoked had higher increases in augmentation index. Changes in smoking behavior were not associated with longitudinal change in other arterial hemodynamic measures. We present the observed (unadjusted) values of central arterial hemodynamic measures between two visits for the longitudinal sample in S3 Table.

**Table 4.** Longitudinal change in central arterial hemodynamic measures between two examination visits by smoking behavior.

| Outcome Variable | Persistently<br>Never Smoked | Persistently<br>Quit | Recently<br>Quit | Persistently<br>Smoking |
| --- | --- | --- | --- | --- |
| Augmentation index, % | 3.45±0.37<br>(reference) | 4.62±0.41<br>(<0.001) | 4.78±0.83<br>(0.09) | 5.48±0.70<br>(0.002) |
| niCFPWV, ms/m | 11.34±0.63<br>(reference) | 11.58±0.69<br>(0.63) | 13.99±1.39<br>(0.04) | 12.87±1.18<br>(0.15) |
| Central pulse pressure, mm Hg | 4.41±0.53<br>(reference) | 5.70±0.59<br>(0.003) | 5.47±1.21<br>(0.35) | 6.00±1.01<br>(0.09) |
| Forward pressure wave, mm Hg | 1.29±0.42<br>(reference) | 1.99±0.46<br>(0.04) | 1.92±0.95<br>(0.49) | 2.37±0.79<br>(0.14) |
| Characteristic impedance, dyne•sec/cm<sup>5</sup> | 14.5±2.6<br>(reference) | 13.9±2.9<br>(0.77) | 12.55±5.87<br>(0.72) | 13.60±4.90<br>(0.84) |
| Global reflection coefficient, % | 0.019±0.002<br>(reference) | 0.021±0.003<br>(0.30) | 0.023±0.005<br>(0.41) | 0.018±0.004<br>(0.77) |
| Peak aortic flow rate, mL/sec | -20.2±1.8<br>(reference) | -17.6±2.0<br>(0.08) | -15.9±4.1<br>(0.26) | -16.8±3.4<br>(0.28) |

niCFPWV, negative inverse carotid-femoral pulse wave velocity. We present least squares means estimates±standard errors (*P*) based on regression models. Participants who formerly quit, recently quit, and were currently smoking are compared to participants who never smoked. Bonferroni-adjusted *P* values ( $P=0.05/21=0.0024$ ) were used to assess significance of associations. Multivariable models adjusted for age, age<sup>2</sup> sex, cohort, body mass index, tonometry heart rate, mean arterial pressure, total to high-density lipoprotein cholesterol ratio, triglycerides, prevalent cardiovascular disease, lipid-lowering medication use, hypertension treatment, and prevalent diabetes, and vascular measure value at the index examination visit. Participants with a change in smoking status of never smoked to quit, never smoked to currently smoking, and quit to currently smoking were excluded from the analysis due to low statistical power to resolve differences.

## Discussion

We investigated the cross-sectional and longitudinal relations of smoking behavior and intensity with measures of central arterial hemodynamics in 2 generations of the community-based Framingham Heart Study. In cross-sectional models, participants who formerly quit and were currently smoking had a higher augmentation index compared to participants who never smoked. Compared to participants who never smoked, participants who currently smoked had higher mean central pulse pressure, global reflection coefficient, and peak aortic flow rate. Higher smoking intensity (cigarettes per day and pack-years) was associated with higher augmentation index and central pulse pressure. The association of smoking cessation (time since quit) with augmentation index, characteristic impedance, and central pulse pressure diminished among participants with longer duration of smoking cessation. Between exam visits, augmentation index increased more among participants who were persistently smoking and who had persistently quit smoking compared to participants who had never smoked. Central arterial hemodynamic measures, particularly indices related to central wave reflection and pressure augmentation, are sensitive to smoking status and intensity and changes in smoking behavior. Longer smoking cessation may partially restore central arterial measures to levels akin to those observed in individuals with lower smoking exposure.

Previous studies examining the acute and chronic effects of smoking on arterial stiffening are controversial and mixed; however, many studies consistently suggest that chronic changes in arterial hemodynamics substantially affect peripheral vessels [20, 31–42]. In a recent Gutenberg Health Cohort Study, researchers observed that smoking status and smoking intensity were positively associated with peripheral arterial stiffness and wave reflection (assessed by digital photoplethysmography and digital plethysmography, respectively) in the community [35]. Our data suggest that higher levels of central wave reflection substantially contribute to the aortic hemodynamic physiologic state in the setting of current, chronic smoking. These findings align with previous research conducted within a young, healthy sample by Mahmud and Feely, who reported higher aortic augmentation index in participants who were actively, chronically smoking compared to non-smoking participants [38, 43]. Additionally, higher smoking intensity was associated with higher central augmentation index indicating a potential dose-response relation of smoking with wave reflection. In healthy, young arteries, there is a progressive increase in arterial stiffness from the highly elastic aorta to the smaller resistance arteries, which creates an impedance mismatch and partial reflection of the forward pressure wave [44, 45]. We had hypothesized that smoking may increase the peripheral arterial stiffness relative to central aortic stiffness, thereby shifting reflecting sites more proximal to the heart. Higher impedance mismatch would increase wave reflection and contribute to higher levels of wave reflection (higher augmentation index and global reflection coefficient), as reflecting sites shift more proximal to the heart among participants who ever smoked compared with participants who never smoked. However, in secondary analyses of muscular and peripheral arterial hemodynamics (S2 Table), we observed lower carotid-brachial pulse wave velocity among participants who were currently smoking compared to participants who never smoked. These results suggest that smoking-related differences in central-to-peripheral arterial impedance gradient were not the underlying factor behind our observations.

Smoking is known to affect vascular function and may contribute to differences and longitudinal changes in central arterial hemodynamics across smoking behaviors. Smoking-induced reactive oxygen species production leads to oxidative stress and contributes to endothelial dysfunction [46, 47]. which impairs vascular tone [31, 32, 48, 49]. A hallmark of endothelial dysfunction is lower nitric oxide bioavailability, a characteristic of healthy individuals who smoke [50]. Endothelial dysfunction impairs regulation of small arteries, which can increase wave reflections from peripheral sites [51]. Our study suggests that among individuals who smoke, wave reflections return to the central aorta more prominently, raising the central pressure augmentation without significantly affecting central aortic stiffness. Kelly et al. showed that aortic augmentation index, but not aortic pulse wave velocity, is sensitive to vasoactive drugs [52], which suggest that dysregulation of vascular tone affects wave reflection independently of aortic stiffness. We observed no differences in aortic stiffness and no to modest differences in peripheral arterial stiffness across smoking behavior groups. It is likely that smoking-associated differences in vascular tone regulation (in the absence of major differences in arterial stiffness) is the major contributor to higher augmentation index [53]. Thus, differential dysregulation of small arterial tone may underlie the relatively higher augmentation index (and its increase over time) in participants who have smoked compared to participants who never smoked. Indeed, multiple studies have shown that vascular endothelium-dependent relaxation [46, 54–64] and downstream microvascular function [64–68] are impaired in individuals who smoke. However, the differences and changes to measures of small and microvascular arterial function due to chronic smoking and changes in smoking behaviors warrants further investigation. In the current study, quitting smoking may gradually reduce the influence of smoking on central wave reflection, indicating that the potential effect of smoking may be partially reversible with longer periods since quitting. However, the impact of past smoking may persist, particularly with a shorter duration since quitting. This concept is underscored in the current longitudinal analysis, which showed that participants who consistently quit smoking still experienced a higher increase in central augmentation index than those who had never smoked, although the increase was less than that of individuals who continued to smoke.

Smoking status and smoking intensity contribute to elevated aortic pressure pulsatility. We observed higher central wave reflection and augmented central pulse pressure among participants who currently smoked compared to participants who never smoked. Similar to augmentation index, the association of smoking with central pulse pressure also demonstrated a dose-response relation. Taken together, higher pressure pulsatility and the earlier arrival of central wave reflection during systole (instead of diastole) may impair coronary perfusion [69, 70]. Higher central pulse pressure increases afterload and may alter cardiac geometry maladaptively as concentric remodeling for individuals who smoke [71–75]. While the traditional view relates elevated central pressure with concentric remodeling due to pressure overload, animal studies have shown that smoking or nicotine exposure contributes to eccentric or mixed cardiovascular remodeling patterns [76–78]. Similar to the current study, Markus et al. observed in two separate cohorts that participants who were currently smoking had higher mean central systolic blood pressure, augmentation index, and left ventricular mass compared to participants who did not smoke [41]. Additionally, in their longitudinal analyses, participants who were currently smoking showed a significant increase in left ventricular mass index, with both an increase in left ventricular wall thickness and end-diastolic diameter; whereas, LV end-diastolic diameter decreased among participants who did not smoke, consistent with aging-associated concentric remodeling [41]. Recently, Fried et al. showed a phenotypic shift from concentric hypertrophy to eccentric hypertrophy in nicotine-exposed, hemodynamically-stressed mice [78]. Thus, higher central aortic pressure and enhanced wave reflection in the setting of smoking may contribute to eccentric remodeling and subsequent decompensation to incident CVD. Our study reveals that quitting smoking may reduce the putative effect of smoking on central pulse pressure in a non-linear pattern, so, similar to wave reflection, the higher hemodynamic burden from smoking might be partially reversed over time. Further studies that assess the mechanisms of smoking and smoking cessation on vascular dysfunction and CVD pathogenesis and prevention of CVD events are warranted.

Despite differences in other central arterial hemodynamic measures, aortic stiffness was similar in participants who never smoked compared to participants who ever smoked in our sample, as indicated by no significant differences in the carotid-femoral pulse wave velocity among smoking behavior groups. This observation is consistent with prior work in assessing differences in chronic smoking [19, 79, 80]. However, our observations are in contrast with other studies that assessed aortic stiffness via MRI or ultrasound methods [40, 81, 82]. For example, in the Multi-Ethnic Study of Atherosclerosis Study, researchers observed higher aortic stiffness (assessed via MRI) among participants who were currently smoking compared to participants who had quit smoking and had never smoked [40]. Assessing aortic stiffness with MRI involves calculating the change in aortic cross-sectional area with pulsatile blood flow and normalizing this by the brachial pulse pressure. This method does not directly measure the pulse wave in the aorta (like arterial tonometry), which may contribute to differences between the MRI investigations and the current study. Additionally, areas of aortic calcification can interfere with the accuracy of MRI measurements, and errors in blood pressure measurements can significantly affect the aortic distensibility calculation, particularly when brachial pressures are used to estimate aortic pulse pressure [83, 84]. Although epidemiologic studies have shown that higher pulse pressure is associated with lower aortic root diameter [85–87], multiple studies show that smoking is associated with higher aortic diameter [86, 88–91].

A recent mechanistic study in mice revealed significant interactions between nicotine exposure and hypertension on aortic diameter and cross-sectional area consistent with eccentric remodeling of the aorta [76]. A transition from concentric to eccentric remodeling may reflect the ongoing adaptation of the aorta to fluctuating and evolving physiological demands and stressors due to smoking. The higher cross-sectional area in individuals who chronically smoke is consistent with our observation of higher peak aortic flow rates in currently smoking participants compared to participants who never smoked. Additionally, we observed a significant non-linear association of time since quitting smoking and characteristic impedance, suggesting a threshold effect after about 25 years. While carotid-femoral pulse wave velocity shows limited sensitivity to changes in aortic diameter, characteristic impedance demonstrates a high degree of sensitivity to variations in the aortic diameter [92]. Considering the inverse relation of characteristic impedance with aortic cross-sectional area, our observations suggest that a relatively longer period of smoking cessation could partially reverse the smoking-induced effects on characteristic impedance. Smoking also increases blood viscosity [93–95], which raises characteristic impedance. However, blood viscosity has limited direct effect on characteristic impedance, which is dominated by inertial rather than viscous effects [96]. Additionally, Shimada et al. reported that blood viscosity was markedly reduced in only a few months after smoking cessation [97]. Given that we observed differences in characteristic impedance only after an extended period of smoking cessation, it appears that the probable recovery of characteristic impedance may be attributed to changes in aortic structure rather than to restoration of blood viscosity. However, future observational and animal experimental research is needed to provide further insight into the effect of smoking and smoking cessation on central arterial hemodynamics and vascular remodeling.

## Limitations

Our study has limitations that should be considered. Although we were able to establish a temporal relation in our longitudinal analyses, the observational nature of the study limits causal inference. The gold standard for establishing causality is randomized, blinded placebo-controlled trials, but studying smoking exposures as a randomized study in humans would be unethical. Additionally, we cannot dismiss the possibility of residual confounding by unknown or unmeasured factors. We used self-reported smoking data, and cotinine levels were not measured in FHS, preventing us from validating smoking behaviors, which may lead to misclassification of participants who underreport or inaccurately recall or report their smoking behaviors. In particular, time since quitting and the amount of smoking may be misclassified. We did not account for e-cigarettes, cigars, and hookah (waterpipe or shisha) or cannabis use; therefore, our study may underestimate the total tobacco and smoking exposure of the participants. In the longitudinal analyses, participants with a change in smoking status of never smoked to quit (n=3), never smoked to currently smoking (n=5), and quit to currently smoking (n=41) were excluded from the analysis due to low statistical power to resolve differences. Although our sample includes participants of the Omni 1 and Omni 2 cohorts from underrepresented racial and ethnic groups, most participants were White individuals of European ancestry; therefore, our findings may not be generalizable to other ethnic or racial groups. Similarly, our sample comprised middle-aged to older participants, so our findings may not be generalizable to younger individuals.

## Conclusion

We examined the cross-sectional and longitudinal relations of smoking behavior and intensity with central arterial hemodynamic measures in middle-aged to older participants of the Framingham Heart Study. Our results suggest that smoking is associated with increases in relative wave reflection, aortic pressure pulsatility, and aortic flow, with associations more pronounced among those who are currently smoking and have smoked compared to those who never smoked, with higher levels among those with greater smoking intensity. However, longer smoking cessation may be associated with reduced relations over time, indicating a potential restoration of vascular function toward levels observed in individuals who never smoked. Relative wave reflection notably was associated with continuing to smoke or quitting compared to those who never smoked after one exam cycle, emphasizing the association of smoking and cessation on central arterial function in a relatively short time. Our study underscores the connection between smoking and cessation on central arterial hemodynamics, highlighting the sensitivity of vascular function to smoking behaviors and intensity.

## Data Availability

All relevant data are within the manuscript and its Supporting Information files.

## Acknowledgements

We thank the Framingham Heart Study of the National Heart Lung and Blood Institute of the National Institutes of Health and Boston University Chobanian and Avedisian School of Medicine. We thank Dr. Kathryn Barger whose expertise in statistical analysis profoundly enhanced the quality of this research.

## Supporting information

**S1 Table. Observed arterial hemodynamic measures stratified by smoking status at the index examination visit.** CFPWV, carotid-femoral pulse wave velocity. niCFPWV, negative inverse carotid-femoral pulse wave velocity. CBPWV, carotid-brachial pulse wave velocity. CRPWV, carotid-radial pulse wave velocity. Values are mean±standard deviation or number (%).

**S2 Table. Estimated least squares means of peripheral arterial hemodynamic measures according to smoking status at Visit 1.** PWV, pulse wave velocity. We present least squares means estimates±standard deviation (*P*) based on regression models. Participants who formerly quit, recently quit, and were currently smoking are compared to participants who never smoked. Bonferroni-adjusted *P* values (*P*=0.05/12=0.0042) were used to assess significance of associations. Multivariable models adjusted for age, age^2^ sex, cohort, body mass index, tonometry heart rate, mean arterial pressure (except where mean arterial pressure is the dependent variable), total to high-density lipoprotein cholesterol ratio, triglycerides, prevalent cardiovascular disease, lipid-lowering medication use, hypertension treatment, and prevalent diabetes.

**S3 Table. Observed central arterial hemodynamic measures between two visits for the longitudinal sample.** CFPWV, carotid-femoral pulse wave velocity. niCFPWV, negative inverse carotid-femoral pulse wave velocity. Values are mean±standard deviation.

## Notes

### Competing Interest Statement

G.F.M. is owner of Cardiovascular Engineering, Inc., a company that designs and manufactures devices that measure vascular stiffness. The company uses these devices in clinical trials that evaluate the effects of diseases and interventions on vascular stiffness. G.F.M. also serves as a consultant to and receives grants and honoraria from Novartis, Merck, Bayer, Servier, Philips, and deCODE genetics. The remaining authors have no disclosures to report.

### Funding Statement

Yes

### Author Declarations

All protocols were approved by the Boston University Medical Center’s Institutional Review Board (https://www.bumc.bu.edu/irb/), and all participants provided written informed consent.

